# Increased ambulance attendances related to suicide and self-injury in response to the pandemic in Australia

**DOI:** 10.1101/2022.05.29.22273082

**Authors:** James John, Ei Phyu Synn, Teresa Winata, Valsamma Eapen, Ping-I Lin

## Abstract

Investigating the change in the trend of ambulance service utilization for suicide in response to the COVID-19 pandemic can clarify the impact of the pandemic on acute health services. To determine whether the trends of the ambulance attendances related to self-injury and suicide changes in response to the COVID-19 pandemic. We extracted the data from the National Ambulance Surveillance System in Australia between March 2018 and March 2021 to examine the trajectory of the ambulance attendances related to self-injury and suicide. The results indicate that the number of ambulance attendances related to self-injury, suicidal ideation and suicidal attempt increased immediately during the first quarter since the outbreak and stayed higher over the at least 12 months. Notably, the post-outbreak surge in ambulance attendances associated with these mental health crises did not continue to escalate further. To sum up, the overall increase in ambulance attendances may reflect increased distress in the community, but also signify disruptions of other non-emergency health services. In contrast to recent evidence for the suicide rate being unchanged during the pandemic, our findings provide a different perspective on the impact of the pandemic on mental health services. This warrants a re-assessment of resources for mental health services in the post-COVID era.

---

Recent evidence indicates that suicide is the leading cause of death among people aged 15–44 years in Australia (Australian Institute of Health and Welfare, 2020). Although there is increasing concerns about the impact of the COVID-19 pandemic on the self-harm and suicide rates, initial evidence in 2020 suggests that the pandemic did not result in increased suicide deaths, contrarily, resulting in a 5.4% drop since the previous year (Clapperton et al., 2021). However, it is unclear whether the impact of the pandemic on suicide based on fatal suicides can reflect how the pandemic affects the capacity of health services.

As the multi-wave course COVID-19 pandemic is being unfolded, the role of surveillance systems in assessing the disease burden has become increasing important (Ogeil et al., 2021). Many surveillance systems for self-harm and suicide using routinely collected information from coroner’s records or hospitalization data might not well represent actual suicide rates due to prolonged time in determining suicide intent (De Leo et al., 2010) and overlooked the cases that were not hospitalized. Therefore, these surveillance approaches only captured a proportion of cases with suicidality and could, hence, under-estimate the magnitude of its burden on the healthcare system.

Ambulance services are often frequented as the frontline health response to acute crises, with evidence showing a substantial amount of paramedic’s time attending to mental health-related presentations including self-harm and suicide-related presentations (Lloyd et al., 2013). Therefore, ambulance attendance (AA) data provides an excellent opportunity to address the above limitations by providing unique information that may not be captured in the hospital or coroner’s data (Lubman et al., 2020). The AA data could also capture patient experiences in self-harm behaviours outside of the inpatient units as data fields and free text (Lubman et al., 2020).

To date, little is known regarding the trends in the self-harm and suicide-related ambulance attendances in context to the pandemic. A recent study in New Zealand showed that the lockdown was associated with a significant increase in AAs for mental health conditions; however, it remains unclear whether AAs related to suicide was also on the rise (Dicker et al., 2020). Therefore, the current study addresses the knowledge gap in examining the patterns of harm and suicide-related AAs associated with the COVID-19 pandemic between March 2018 and March 2021, in Australia.

The overall aim of this study is to determine the trends in ambulance attendances for self-harm, suicidal ideation, and suicide attempts associated with the COVID-19 pandemic between March 2018 and March 2021. We hypothesize that increase in the ambulance attendances for self-harm, suicidal ideation, and suicide attempts will be associated with time since COVID-19 (March 2020 onwards) compared to the time before the pandemic.

To examine the pandemic’s impact, we extracted 1-month per quarter data on AAs related to self-injury, suicidal ideation, and suicide attempts in Australia during March 2018 - March 2021 from the National Ambulance Surveillance System (NASS) (Lubman et al., 2020). March 2020 was considered the cutoff between the pre-pandemic and COVID-19 periods.

Independent samples t-tests were used to compare mean differences in the reported number of self-injuries, suicidal ideation, and suicide attempts between pre-COVID and post-COVID phases at both the state and national levels. Interrupted Time Series (ITS) analysis was used to determine whether the trajectories of self-injury, suicidal ideation, and suicide attempts related AAs in Australia between March 2018 and March 2021 substantially change in response to the COVID-19 pandemic that hit Australia around January 2020 on top of the time effect. The ITS regression model with two interventions takes the following form:

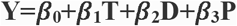

Where Y denotes the outcome variable; T is the time since the start of the sample; D is a dummy variable representing the first COVID-19 period (March 2020, pre-COVID period 0, otherwise 1); P is the time since the first COVID-19 quarter.

In our regression model, β_0_ represents the baseline level of the dependent variable; β_1_ is the slope of the line before the first COVID-19 period; β_2_ measures the immediate effect that occurs after the first COVID-19 period; β_3_ represents the difference of the slope between pre- and post-outbreak periods - which represents the sustained effect of the COVID-19 pandemic.

The scatter plots for rates of self-injury, suicidal ideation, and suicide attempts were created to visually inspect our data and examine whether COVID-19 had a significant impact or not. The observed values of the rates for self-injury, suicidal ideation, and suicide attempt, compared with their corresponding counterfactual values (i.e., events had the pandemic not occurred) were evaluated. Delayed effect (6 months after first COVID-19 period) was also assessed if there was no immediate effect at the first period. Autocorrelation was examined using the Durbin-Watson test with the significance threshold at p < 0.05. All analyses were conducted using the R software (Version R 4.1. 2 “Bird Hippie”).

The average number of ambulance attendances related to self-injuries, suicidal ideation, and suicide attempts related, overall (all four states combined) and stratified by four states with the mean differences before and since the COVID-19 pandemic are presented in Table 1.

**Table 1.**
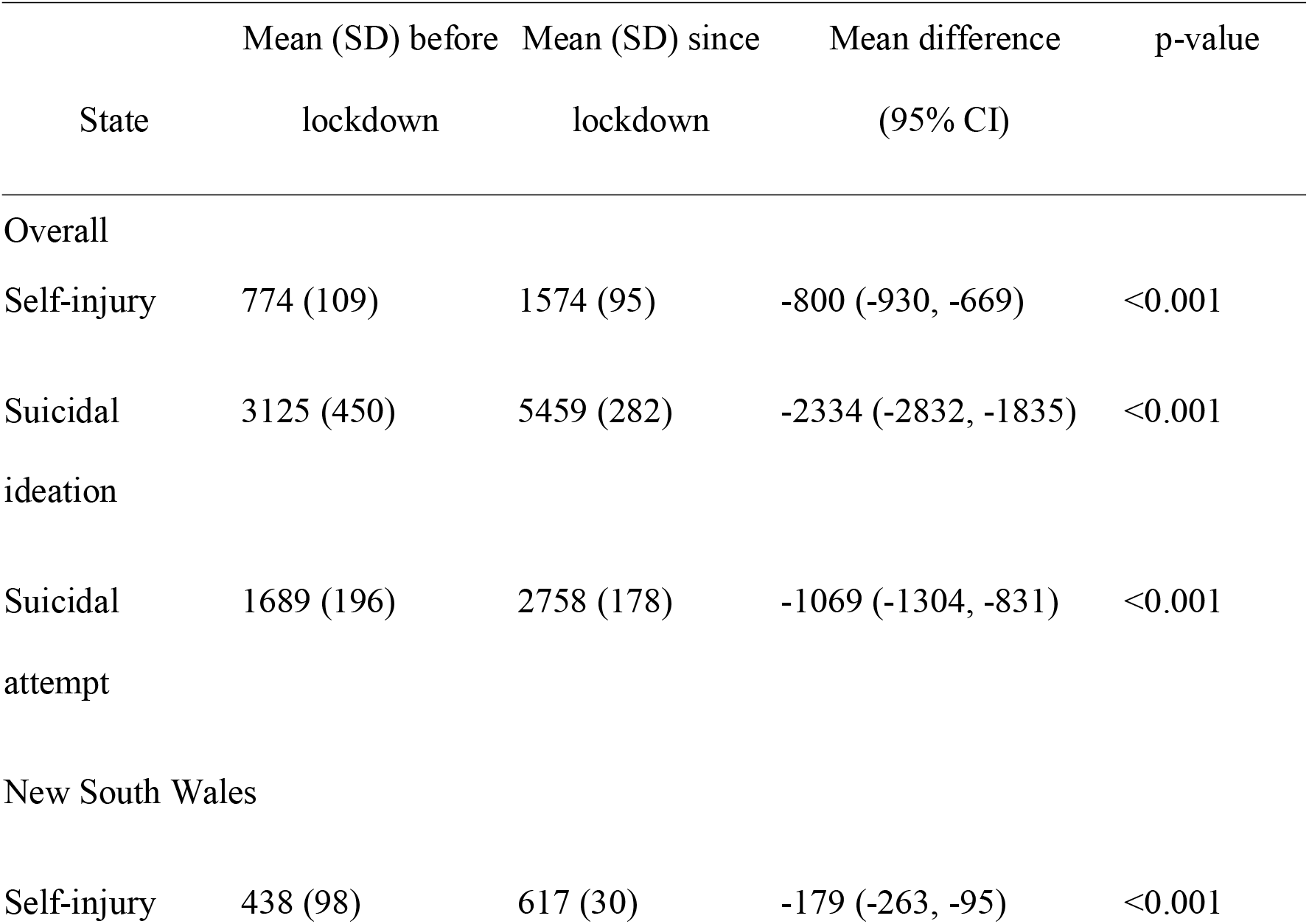
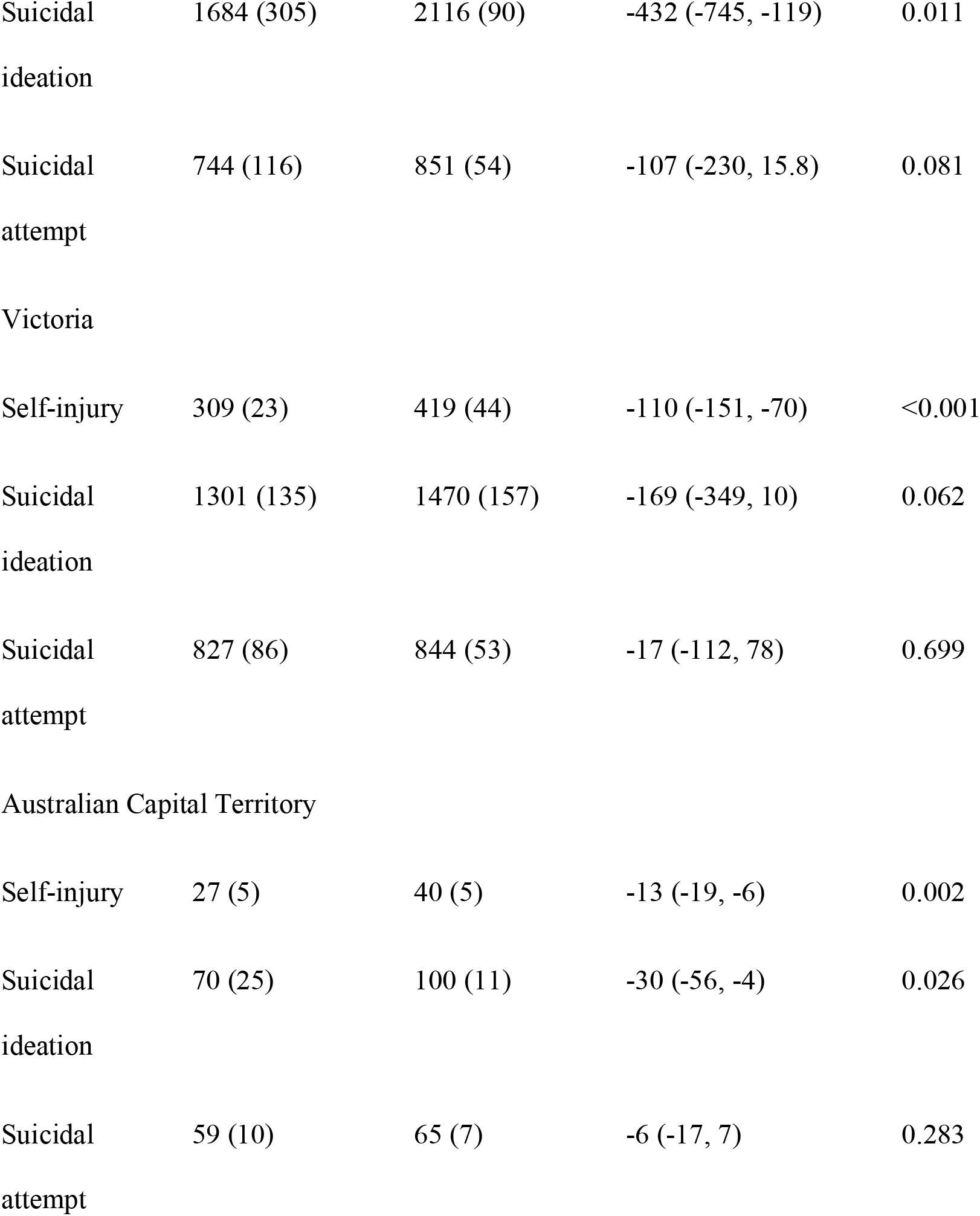
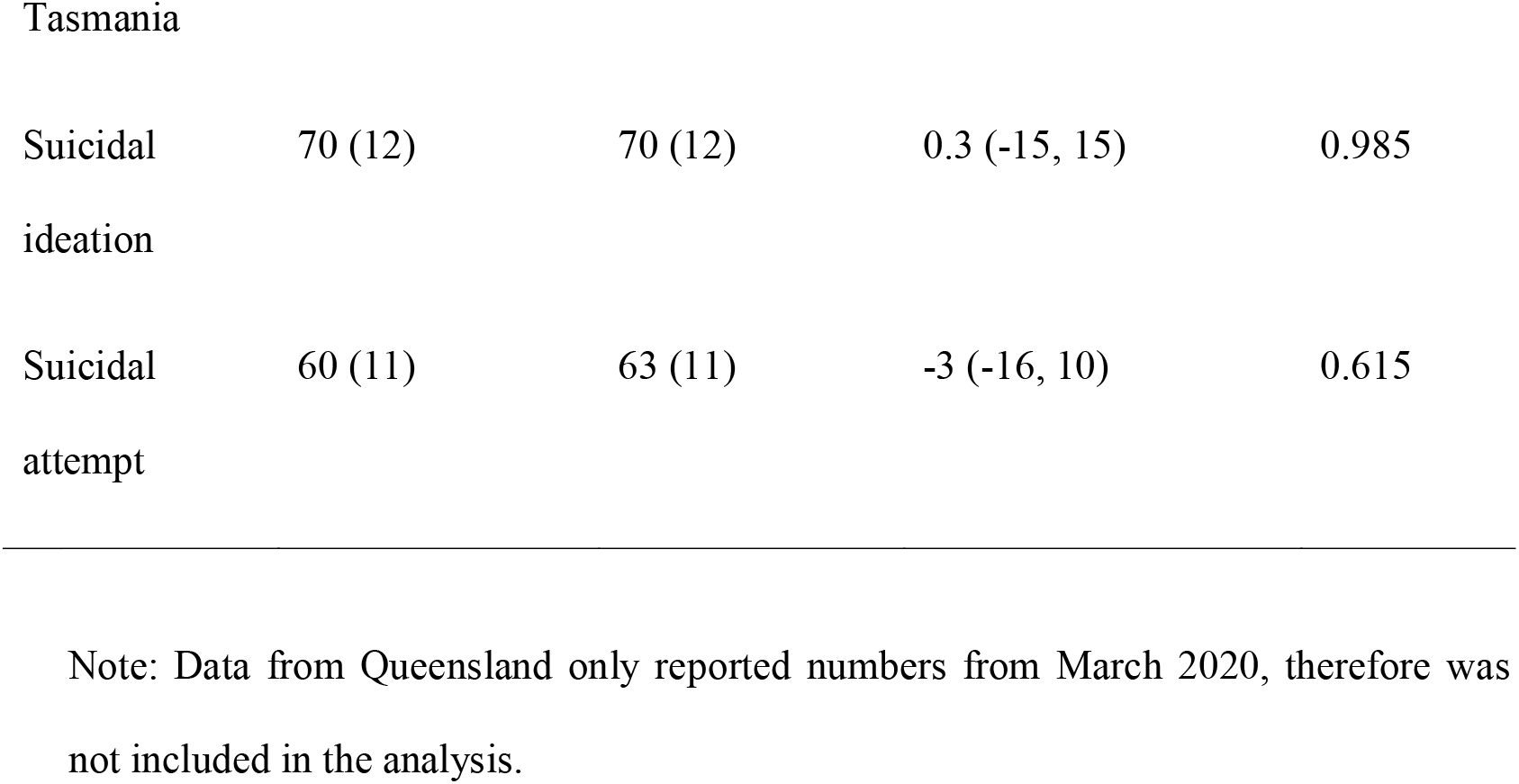
Ambulance attendances related to self-injuries, suicidal ideation, and suicide attempts before and since the COVID-19 pandemic.

We have presented only the key findings of the ITS analysis in Table 2. Overall, the trajectories of AA for the numbers of self-injury, suicidal ideation, and suicide attempts, increased substantially (p-value <0.05) during the first wave immediately (β_2_) after the outbreak of COVID-19. However, the sustained effect (β_3_) did not differ significantly between pre- and post-outbreak periods (Figure 1). The abrupt increment in ambulance attendances reflected the difference between March 2020 and December 2019, which did not imply that the increase began as early as December 2019. In fact, the first case of COVID-19 was documented on 25 January 2020.

**Table 2.**
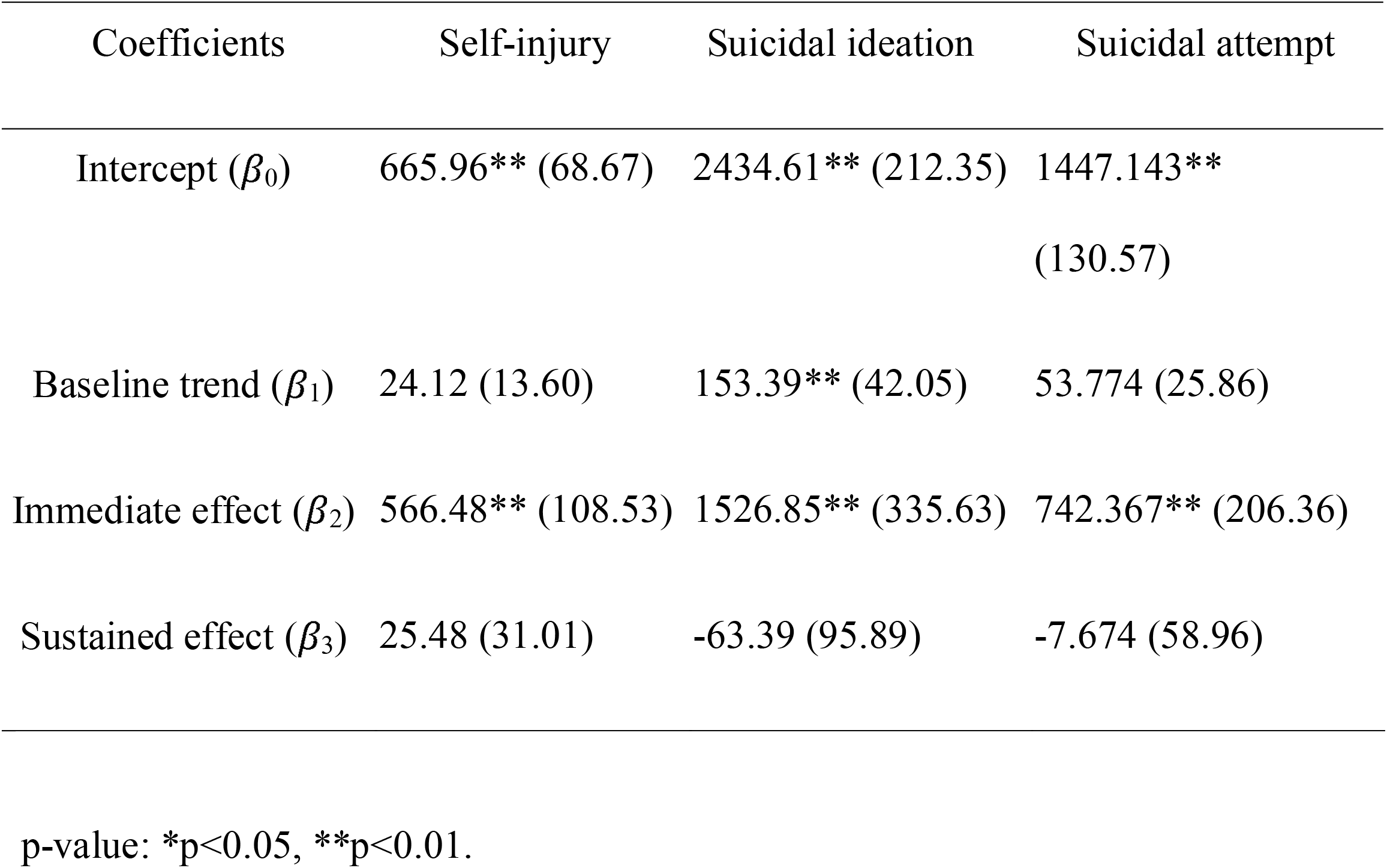
ITS analysis of overall self-injury, suicidal ideation, and suicide attempts related AAs.

**Figure 1.**
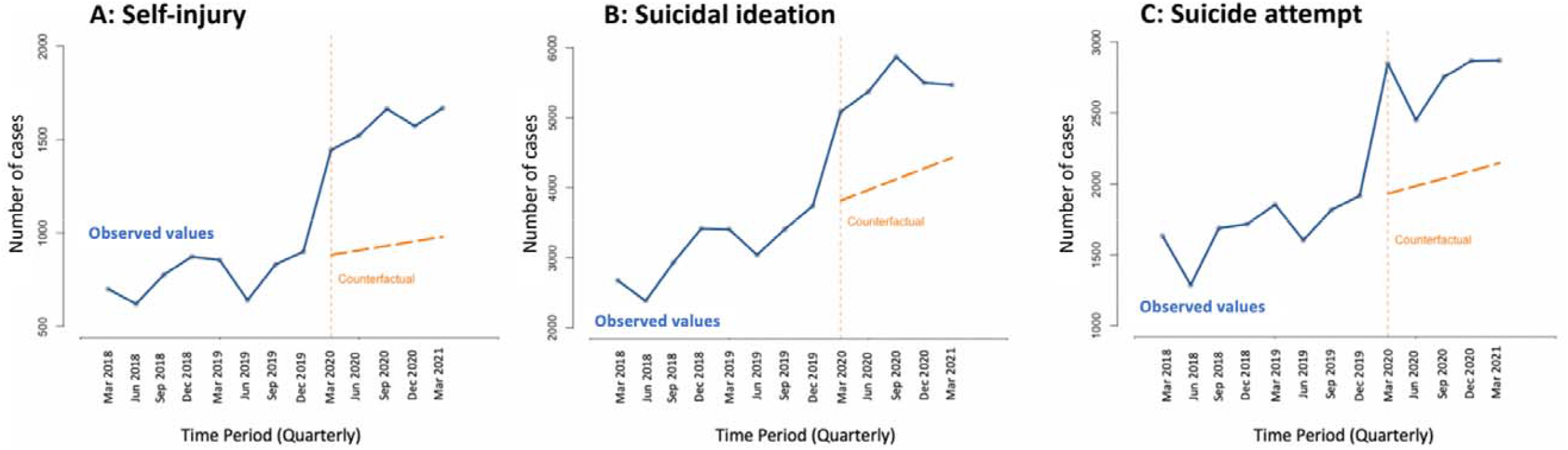
Trends of ambulance attendances (AAs) related to self-injury, suicidal ideation, and suicide attempts. Observed values - blue line; counterfactual – orange dotted line; Vertical dotted line at March 2020 indicates COVID-19 period.

We found the AAs related to self-injury or suicide increased substantially during the first wave of COVID-19 in Australia compared to pre-pandemic levels. Although there was a lack of a further escalation in the 12 months following the outbreak, the increase did not subside in that same post-outbreak period, either. This may not be entirely attributable to increased distress in the population since some of the cases warranting clinical attention might turn to ambulance services due to interruption in other clinical services. Our study is limited with the use of 1-month per quarter snapshots of case numbers which may not be representative of total attendances in a quarter and also does not account for multiple attendances for the same patient.

Our findings provide a different perspective from a previous international study indicating a unchanged risk of fatal suicide during the pandemic (Pirkis et al., 2021). Focusing on the data on mortalities or hospitalization records may be biased towards more serious cases that need more intensive clinical care (Williams, 2015), which could under-estimate the impact of the pandemic on this mental health crisis. Our study, hence, suggests a critical need to re-evaluate the impact of the pandemic on mental health crises and the capacity of clinical services.

## Data Availability

All data produced are available online at

https://www.aihw.gov.au/suicide-self-harm-monitoring/data/data-downloads

## Ethics approval

The study involves anonymous aggregate data freely available in the public domain and does not require ethics approval.

